# Investigating the Impact of Sex on Outcomes in Juvenile Idiopathic Arthritis

**DOI:** 10.64898/2026.03.24.26349201

**Authors:** Wong Sa, Stephanie Shoop-Worrall, Gavin Cleary, Flora McErlane, Lucy Wedderburn, Kimme L Hyrich, Coziana Ciurtin

## Abstract

**Background:** Juvenile idiopathic arthritis (JIA) shows recognised sex differences, but their impact on treatment and early outcomes remains uncertain. This study assesses sex-specific patterns in onset, phenotype, treatment timing, and short- and medium-term outcomes in JIA.

**Methods:** Data were drawn from the Childhood Arthritis Prospective Study (CAPS), a UK multicentre inception cohort of 1,789 children presenting with a new episode of arthritis. Demographics, subtype distribution, clinical features, and 6- and 12-month outcomes were stratified by sex. Cox, Kaplan-Meier, and linear regression models assessed associations between sex and treatment initiation and 12-month outcomes.

**Results:** The cohort was predominantly female (64.3%), with a median age at symptom onset of 6.8 years. Girls were younger than boys at onset (6.1 vs 7.8 years, p<0.0001) and diagnosis (7.0 vs 9.1 years, p<0.0001) and demonstrated a distinct bimodal age distribution. Diagnostic delay was short and comparable (median 4.4 months, p=0.8932). At diagnosis, girls had slightly higher active joint counts (p=0.0080, while inflammatory markers were similar except in psoriatic JIA, where females had higher ESR and CRP. After adjustment, sex was not associated with time to methotrexate (HR 0.89, 95% CI 0.74-1.06) or biologic initiation (HR 0.91, 95% CI 0.72-1.16). Outcomes at 6- and 12-month were largely comparable, with only ESR showing a modest male-favoured improvement at 12 months (p=0.0480).

**Conclusions:** Sex shaped age at onset and subtype distribution but did not independently influence treatment timing or early outcomes, underscoring the value of sex-aware analyses while confirming broadly comparable short-term trajectories in JIA.

*Evidence before this study:* Recent evidence on sex effects in JIA is genuinely mixed: several cohorts have reported that girls, despite more severe onset, show greater resolution of objective inflammation, while a newer, large network analysis found females had poorer outcomes across composite disease activity and pain, pointing to potential inequities or phenotype-driven differences. In parallel, boys, especially in enthesitis-related arthritis (ERA), often exhibit more persistent activity and risk of damage. Overall, the picture is controversial: sex appears to shape biology, trajectory, and patient-reported burden in different ways across subtypes and settings, reinforcing the need for sex-stratified analyses, careful adjustment for confounders, and precision approaches that integrate biomarkers, subtype, and social context in JIA care.

*Added value of this study:* The study establishes that, although sex is closely linked to JIA subtype distribution and baseline clinical features, it does not independently determine the timing of methotrexate or biologic initiation within a UK inception cohort. By analysing one of Europe’s largest prospective multicentre datasets, it provides strong evidence that treatment decisions appear to be guided by disease characteristics rather than demographic bias. Within the context of the UK National Health Service (NHS), where universal access to paediatric rheumatology care is a core principle, this study provides important epidemiological evidence on sex and equity in JIA. Although clear sex differences were observed in age at onset, subtype distribution, and certain diagnostic features, these did not translate into disparities in treatment timing or medium-term disease burden. The absence of sex-based differences in 6 and 12-month outcomes, despite variation in baseline presentation, suggests that the NHS model of coordinated, guideline-driven care may help buffer against inequities that might otherwise emerge in systems with variable access. These findings reinforce the value of population-based cohorts in evaluating equity within healthcare delivery and highlight that, in this setting, sex does not appear to drive differential treatment or short-term clinical trajectories.

*Implications of all the available evidence.:* This study underscores sex as an important biological variable in JIA. Although treatment initiation was equitable and disease-driven, baseline phenotype differences and isolated effects on 12-month outcomes highlight how sex interacts with JIA subtype and initial disease burden. Prior work shows that females often present earlier with higher inflammatory burden, while males are more frequently affected by ERA, a subtype linked to treatment resistance and poorer long-term outcomes. Yet published findings remain inconsistent, with some cohorts reporting better resolution of inflammation in females and others suggesting poorer outcomes. Our findings suggest that coordinated and guideline-driven care may minimise sex-related disparities in JIA, even when underlying biological or phenotypic differences exist. The comparable medium-term trajectories observed across sexes support equitable paediatric rheumatology care in the UK and highlight the need to continue monitoring for structural or access-related inequities beyond clinical measures.

## Introduction

Juvenile idiopathic arthritis (JIA) is the most prevalent chronic rheumatic condition in children. Around 12,000 children and young people (CYP) in the UK are affected by JIA, making it one of the most common causes of chronic physical disability in childhood (1). A recent national study using electronic health records estimated the annual incidence of JIA in England at approximately 5 to 10 per 100,000 CYP, with variations across ethnic groups (2). Sex bias in JIA is a well-documented phenomenon, with epidemiological data consistently demonstrating a higher incidence and prevalence in females compared to males, typically with a female-to-male ratio ranging from 2:1 to 3:1 (3). This disparity is most pronounced in subtypes such as oligoarthritis and rheumatoid factor (RF)-negative polyarthritis, whereas enthesitis-related arthritis (ERA) shows a male predominance.

Despite advances in JIA treatment, major gaps remain in tailoring care to each child’s disease course, highlighting the need to move from uniform protocols toward precision, person-centred approaches (4). CYP with JIA need timely, equitable, and holistic management, as well as access to advanced therapies, and coordinated transitional care (5). Even if standards of care for JIA are well established (6), a recent national report showed that CYP in the UK often face diagnostic delays and fragmented care due to limited clinical awareness, unclear referral pathways, and poor coordination between specialist services (7).

Personalised JIA care requires sex-specific analyses, as sex influences JIA onset, severity, treatment response, and lived experience. Yet current guidelines do not stratify recommendations by sex, reflecting limited evidence that sex-related differences warrant distinct therapeutic approaches (8). Sex-specific factors may shape challenges across the JIA life course, but remain under-studied.

Prognosis varies widely and is driven by demographic, clinical, and serological predictors, with JIA subtype-specific factors central to risk stratification. Persistent oligoarthritis and systemic JIA generally show more favourable remission profiles (9). In childhood, girls, who are more frequently affected by oligoarticular and polyarticular JIA subtypes, may face earlier onset and more persistent joint inflammation, impacting physical development and school participation (10, 11). Younger age and male sex are associated with higher remission rates in oligoarthritis, while older age at diagnosis, particularly in females, correlated with better outcomes in RF-negative polyarthritis (12). In contrast, RF-positive polyarthritis and ERA are linked to poorer prognoses, characterised by lower remission rates and greater long-term disease burden (13, 14). RF-positive polyarthritis, in particular, is noted for its severe functional impairment and minimal remission off medication (15). While oligoarticular JIA often has a favourable trajectory, early-onset disease in girls, with persistent activity is a strong predictor of more aggressive joint damage and sustained disability, with initial functional impairment serving as a key indicator of long-term outcomes (12, 16).

In some JIA cohorts, clinical outcomes and disease severity overall seem to be influenced by sex, with some studies reporting more aggressive disease courses and poorer long-term functional outcomes in females (11). In the ReACCh-Out cohort, females with JIA had higher rates of persistent disease with impact on treatment responses (17), as female sex was independently associated with a lower probability of achieving early remission (within 6 months) when treated with NSAIDs and methotrexate (18). In both the Canadian and Nordic JIA cohorts, girls were less likely to reach remission within 12 months, even after adjusting for other clinical variables (19).

In a UK-wide multicentre prospective longitudinal study, girls reported higher pain and fatigue over time, with distinct disease trajectories compared to boys (3). Hormonal changes during puberty can exacerbate disease activity, particularly in females (20), while psychosocial stressors like body image and peer dynamics may compound the sex-biased disease burden (21). In adulthood, women with a history of JIA often report higher rates of chronic pain, fatigue, and reduced quality of life than men with JIA (22).

Sex-based and global disparities in JIA arise from intertwined biological, environmental, and healthcare factors. Worldwide, unequal access to early diagnosis, biologics, and specialist care worsens outcomes, particularly in resource-limited settings, where girls may also face additional barriers linked to gender norms and structural inequities (23). Addressing these gaps requires sex-aware research and equitable, culturally responsive care.

Despite established sex differences in JIA presentation, examining sex as an outcome remains essential, as sex-linked genetic, hormonal, immunological, and social factors may shape disease course and treatment response. This study evaluates how sex influences age at onset, subtype distribution, serology, diagnostic delay, and timing of methotrexate and biologic initiation, and how these relate to outcomes at 6 and 12 months in a UK-wide inception cohort. The aim is to determine whether sex-based disparities persist within the NHS, where all children receive free care at the point of use.

## Methods

This study used data from the Childhood Arthritis Prospective Study (CAPS), a UK multicentre longitudinal inception cohort established in 2001 and approved by the UK Northwest Multicentre Research Ethics Committee (MREC 00/8/53). Children and young people with new-onset arthritis before age 16, diagnosed as JIA by ILAR criteria (24), were included. Data were collected at baseline, 6 months, and 12 months. Variables analysed included demographics (sex, ethnicity, socioeconomic status), clinical history (age at symptom onset, diagnostic interval, breastfeeding, family history), and all six JIA core outcome measures, including joint counts, global and functional assessments, inflammatory markers, and autoantibodies. Treatment timelines, including time from baseline to DMARD initiation, were also recorded Sex was defined by biological designation at birth and information extracted from medical records or self-reported. Statistical analyses were conducted in R (v4.4.3). Normality and age distribution patterns were assessed visually (Kernel density estimation) and using Shapiro-Wilk tests. Descriptive statistics stratified by sex used appropriate metrics applied based on data distribution. Univariable differences in demographic and clinical characteristics, as well as inflammatory markers between different ILAR subtypes disaggregated by sex were assessed via Chi-squared testing for categorical variables, and Mann-Whitney U-tests for continuous variables. We performed a subgroup analysis to identify sex differences in inflammatory markers across each JIA subtype. Longitudinal analyses included Kaplan-Meier curves and Cox proportional hazards models to evaluate sex differences in time from symptoms onset to methotrexate or biologic treatment initiation, adjusting for age, ILAR subtype, joint count at baseline, and presence of uveitis. Changes in disease activity at 12 months assessments between sexes were analysed using univariable and adjusted multivariable linear regression models.

## Results

### Cohort characteristics

In this study, a cohort of 1,789 CYP with inflammatory arthritis was comprehensively characterised. Baseline characteristics, stratified by sex, are summarised in **Table 1**, alongside data availability across the dataset. The cohort was predominantly female (64.3%; female-to-male ratio 1.8:1) and largely of White ethnicity (89.9%). The median age at symptom onset was 6.8 years [IQR 2.9 - 10.9].

**Table 1:** Baseline characteristics of the JIA cohort stratified by sex.

| **Variable** | **Data availability, n (%)** | **n (%) or median (IQR)** | **Female (n = 952, 64.3)** | **Male (n = 528, 35.7)** | **P-value** |
| --- | --- | --- | --- | --- | --- |
| **Ethnicity** | 1585 (88.6) |  |  |  | 0.554 |
| White |  | 1419 (89.5) | 773 (89.8) | 428 (89.5) |  |
| Asian* |  | 83 (5.24) | 45 (5.23) | 23 (4.81) |  |
| Black |  | 22 (1.39) | 10 (1.16) | 9 (1.88) |  |
| Other** |  | 56 (3.53) | 33 (3.83) | 18 (3.77) |  |
| **Age at baseline (diagnosis) (years)** | 1458 (81.5) | 7.80 (3.6, 12.0) | 7.03 (2.98, 11.7) | 9.08 (5.26, 12.2) | **<0.0001** |
| **Delay from symptoms onset to diagnosis (months)** | 1160 (64.8) | 4.43 (2.1, 8.3) | 4.43 (2.14, 8.02) | 4.43 (2.10, 8.97) | 0.893 |
| **Age at menarche (years)** | 428 (45.0) | 8 (8, 11) | 8 (8, 12) | NA | NA |
| **Vaccines up to date** | 1560 (87.2) | 1464 (93.8) | 793 (93.8) | 427 (92.8) | 0.551 |
| **Number of full siblings** | 1100 (61.5) | 1 (0, 2) | 1 (0, 1) | 1 (1, 2) | **0.0006** |
| **Number of half siblings** | 1075 (60.1) | 0 (0, 0) | 0 (0, 0) | 0 (0, 0) | 0.236 |
| **Breastfed** | 1128 (63.1) | 619 (54.9) | 393 (54.4) | 213 (55.9) | 0.686 |
| **Family history** |  |  |  |  |  |
| Psoriasis | 700 (39.1) | 85 (12.1) | 34 (12.1) | 23 (14.6) | 0.55 |
| HLA-B27+ associated arthritis | 694 (38.8) | 84 (12.1) | 38 (13.3) | 22 (14) | 0.957 |
| IA in 1<sup>st</sup> degree relatives | 197 (11.0) | 17 (8.6) | 11 (9.7) | 4 (5.5) | 0.411 |
| IA in 2nd degree relatives | 197 (11.0) | 54 (27.4) | 33 (29.2) | 20 (27.4) | 0.92 |
| **Source of referral** | 1718 (96.0) |  |  |  | 0.5149 |
| Paediatrician |  | 735 (42.8) | 406 (42.7) | 207 (39.1) |  |
| Orthopaedic surgeon |  | 408 (23.7) | 209 (22.0) | 138 (26.1) |  |
| A & E |  | 121 (7.0) | 69(7.3) | 32 (6.0) |  |
| GP |  | 287 (16.7) | 163 (17.1) | 90 (17.0) |  |
| Physiotherapist |  | 28 (1.6) | 11 (1.2) | 9 (1.7) |  |
| Other | 184 (10.3) | 139 (8.1) | 71 (7.5) | 43 (8.1) |  |
| **Primary carer information\*\*\*** |  |  |  |  |  |
| Age when left education if no degree (years) | 635 (35.5) | 16 (16, 18) | 16 (16, 18) | 16 (16, 18) | 0.796 |
| Completed sixth form | 609 (34.0) | 164 (24.5) | 113 (36.6) | 51 (33.6) | 0.563 |
| Attended college or university | 627 (35.1) | 357 (53.3) | 235 (57) | 122 (59.2) | 0.666 |
| Has degree | 623 (34.8) | 149 (22.2) | 93 (22.7) | 56 (27.5) | 0.231 |
| **1<sup>st</sup> Carer income** | 511 (28.6) |  |  |  | 0.609 |
| <£10k |  | 97 (19.0) | 68 (20.12) | 28 (16.97) |  |
| £10k-£14,999 |  | 95 (18.6) | 60 (17.75) | 32 (19.39) |  |
| £15k-£19,999 |  | 78 (15.3) | 50 (14.80) | 28 (16.97) |  |
| £20k-£24,999 |  | 88 (17.2) | 59 (17.46) | 28 (16.97) |  |
| £25k-£29,999 |  | 45 (8.8) | 33 (9.76) | 12 (7.27) |  |
| £30k-£34,999 |  | 19 (3.7) | 16 (4.73) | 2 (1.21) |  |
| £35k-£39,999 |  | 25 (4.9) | 14 (4.14) | 11 (6.67) |  |
| £>=40k |  | 64 (12.5) | 38 (11.24) | 24 (14.55) |  |

*Legend:* \*Indian, Pakistani, Bangladeshi, Chinese; \*\*Includes mixed race. \*\*\*questions about education level are independent: e.g. carers may answer one and skip another. Carers can attend college without completing sixth form (e.g., vocational routes), or complete sixth form but not attend university or attend university but not complete a degree. A&E – Accident and Emergency; GP – general practitioner.

Girls were significantly younger than boys at both symptom onset (median = 6.1 [IQR 2.4-10.7] vs. 7.8 [IQR 4.5-11.2] years, p<0.0001) and diagnosis (median 7.0 [IQR 3.0 - 11.7] vs. 9.1 [IQR 5.3 - 12.2] years, p<0.0001). Kernel density estimation demonstrated a clear multimodal distribution of age at symptom onset in the overall cohort, with distinct peaks in early childhood (2.1 years) and adolescence (11.4 years) (**Suppl. Figure 1 A**). Sex-stratified analysis revealed that this pattern was primarily driven by females, who exhibited a well-defined bimodal distribution closely mirroring the overall cohort (early-onset: 2.1 years; adolescent-onset: 11.7 years) (**Suppl. Figure 1B**). In contrast, males showed a broader and less distinct distribution, with peaks occurring in mid-childhood (6.1 years) and adolescence (11 years) (**Suppl. Figure 1C**). A near-identical pattern was observed for age at diagnosis, with the overall distribution being multimodal and the most prominent peaks occurring at 2.5 and 12.5 years (**Suppl. Figure 2A**). Females again demonstrated a distinct bimodal distribution, with peaks at 2.6 and 12.5 years (**Suppl. Figure 2B**). In contrast, the male distribution, although non-normal and containing model-identified peaks at 6.8 and 11.9 years, appeared more uniform between these points and lacked the clear separation of subpopulations evident in females (**Suppl. Figure 2C**).

The overall symptom-to-diagnosis interval demonstrated a right-skewed distribution (**Suppl. Figure 3A**), with most receiving a diagnosis within 4.4 months of symptom onset. There were no sex differences in diagnostic delay (median = 4.4 [IQR 2.1 - 8.0) vs. 4.4 [IQR 2.1 - 8.9] months in girls vs. boys, p= 0.893). **(Table 1, Suppl. Figure 3 B-C).** The number of full siblings was lower in girls vs. boys with JIA (median = 1 [IQR 0 - 1] vs. 1 [IQR 1 - 2], p=0.0006), although there were no sex differences in the proportion of CYP with JIA and positive family history of any type of inflammatory arthritis, including HLA-B27 associated arthritis, or psoriasis (**Table 1**).

Most CYP were referred to paediatric rheumatology by general paediatricians (735, 42.8%) or orthopaedic surgeons (408, 23.7%). Referral patterns did not differ by sex (p = 0.5149). Similarly, no sex-based differences were observed across any categories of primary carer education or household income (**Table 1**).

### Impact of sex on JIA phenotype, and clinical and serological markers at baseline

The distribution of JIA subtypes differed significantly by sex (p < 0.001) (**Table 2**). Females predominated in all subtypes except ERA, which was overwhelmingly male (86.2%). The strongest female predominance was observed in the RF-positive polyarticular subtype, with a female-to-male ratio of 5.8:1, whereas ERA demonstrated the opposite pattern, with a male-to-female ratio of 6.2:1 (**Table 2, Figure 1**).

**Figure 1:**
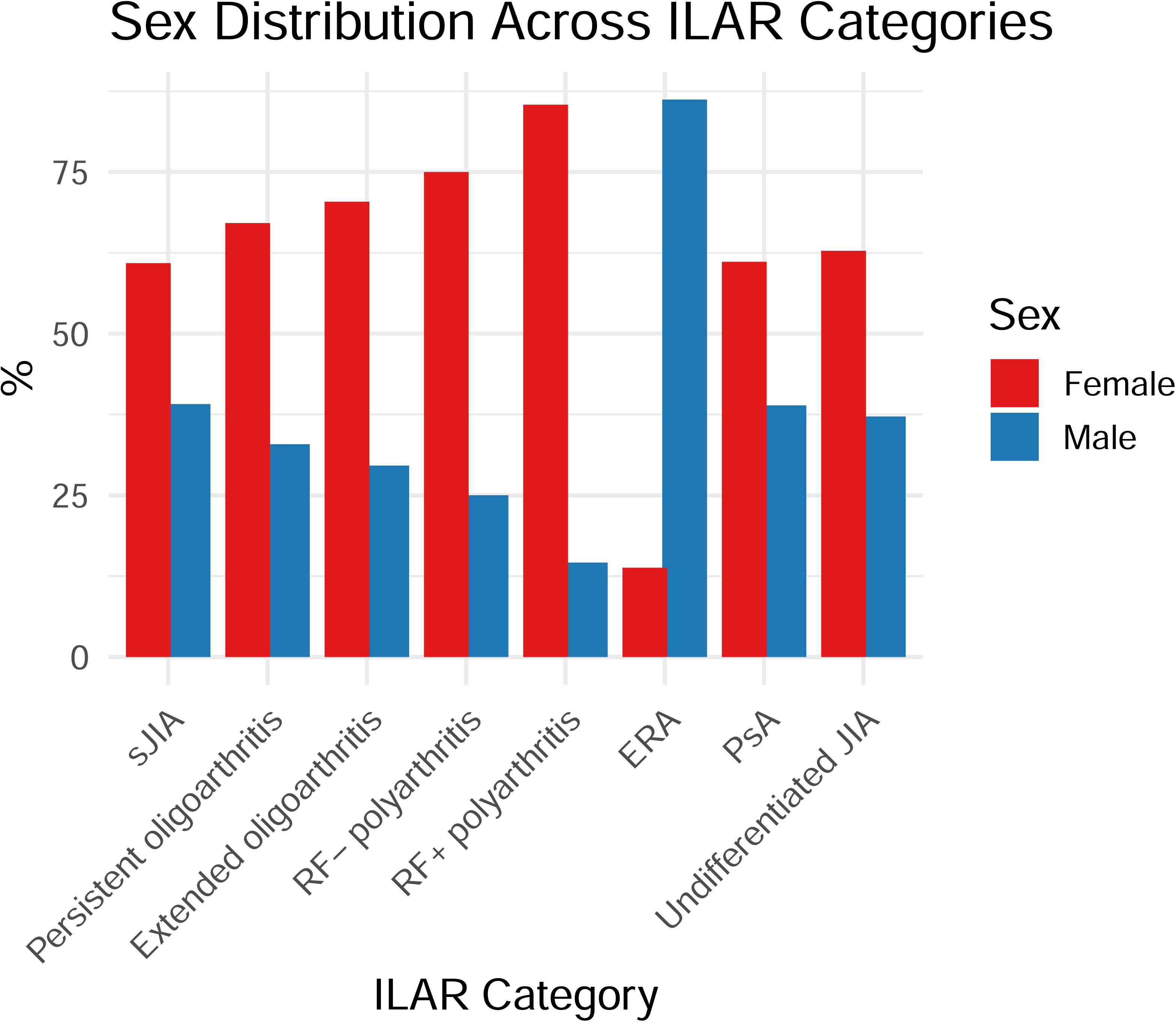
ILAR JIA categories at baseline stratified by sex. *Legend:* ERA - enthesitis-related arthritis; JIA - juvenile idiopathic arthritis; PsA - psoriatic arthritis; RF- polyarthritis - rheumatoid factor-negative polyarticular JIA; RF+ polyarthritis - rheumatoid factor-positive polyarticular JIA; sJIA - systemic onset JIA (Still’s disease).

**Table 2:** JIA phenotype, clinical and serological markers, and patient/parent reported outcomes at diagnosis (baseline) stratified by sex.

|  | n (%) or median (IQR) | Female (n = 952, 64.3%) | Male (n = 528, 35.7%) | P-value |
| --- | --- | --- | --- | --- |
| **ILAR category at baseline (Data available for n=1601, 89.5%)** |  |  |  | **<0.0001** |
| sJIA (Still's) | 95 (5.93) | 53 (5.6) | 34 (6.49) |  |
| Persistent oligoarthritis | 671 (41.9) | 416 (44.1) | 204 (38.9) |  |
| Extended oligoarthritis | 33 (2.06) | 19 (2.0) | 8 (1.53) |  |
| RF -ve polyJIA | 246 (15.3) | 162 (17.2) | 54 (10.3) |  |
| RF +ve polyJIA | 50 (3.12) | 35 (3.7) | 6 (1.15) |  |
| ERA | 76 (4.75) | 9 (1.0) | 56 (10.7) |  |
| Psoriatic arthritis | 97 (6.06) | 55 (5.8) | 35 (6.68) |  |
| Unclassifiable JIA | 277 (17.3) | 169 (17.9) | 100 (19.1) |  |
| Other type of IA | 56 (3.5) | 26 (2.8) | 27 (5.15) |  |
| **Baseline JIA characteristics: n (%) of CYP with available data** |  |  |  |  |
| Active joint count<br>1693 (94.6) | 2 (1, 5) | 2 (1, 5) | 1 (1, 4) | **< 0.0080** |
| Limited joint count<br>1694 (94.7) | 1 (1, 3) | 1 (1,4) | 1 (0, 3) | 0.084 |
| ESR<br>904 (50.5) | 22 (8, 50.3) | 25 (9, 52) | 25 (6, 50) | 0.254 |
| CRP<br>858 (48.0) | 7 (4, 24) | 7.5 (4, 28.4) | 7.4 (4, 25.4) | 0.775 |
| Uveitis<br>1455 (81.3) | 71 (4.9%) | 41 (5.3%) | 21 (5%) | 0.916 |
| Physician GA*<br>1084 (60.6) | 28.5 (15, 50) | 29 (16, 50) | 28 (15, 50) | 0.462 |
| **Patient/parent reported outcomes at baseline** |  |  |  |  |
| CHAQ<br>1135 (63.4) | 0.75 (0.25, 1.375) | 0.75 (0.25, 1.38) | 0.75 (0.125, 1.38) | 0.146 |
| Patient/parent GA*<br>1132 (63.3) | 23 (6, 50) | 21 (5, 49.3) | 25 (5, 50.5) | 0.391 |
| Pain*<br>1058 (59.1) | 30 (8, 60) | 28 (8.75, 57) | 31.5 (7.75, 60) | 0.742 |

Legend: ANA – antinuclear antibodies; ERA – enthesitis related arthritis; GA – global assessment; JIA – juvenile idiopathic arthritis; IA – inflammatory arthritis; ILAR – International League of Associations for Rheumatology; RF -ve poly JIA – rheumatoid factor negative polyarticular JIA; RF +ve – rheumatoid factor positive polyarticular JIA; sJIA – systemic onset juvenile idiopathic arthritis, also known as Still’s disease. \*measured on a visual analogue (VAS) scale 1-10.

At diagnosis, females had higher active joint counts than males (median 2 [IQR 1-5] vs 1 [IQR 1-4], p< 0.0080), while limited joint counts were comparable between the sexes (median 1 [IQR 1 - 4] vs 1 [IQR 0 - 3], p = 0.846). There were no sex-based differences in the prevalence of JIA-associated uveitis (41/770 [5.3%] in girls vs 21/423 [5.0%] in boys, p = 0.916).

Similarly, across all JIA phenotypes, no sex differences were identified in serum inflammatory markers at presentation. Erythrocyte sedimentation rate (ESR) values were comparable between boys and girls (25 [IQR 9-52] vs 25 [IQR 6-50] mm/hr, p = 0.254), as were C-reactive protein (CRP) levels (7.5 [IQR 4-28.4] vs 7.4 [IQR 4-25.4] mg/L, p = 0.775) (**Table 2**).

Sex-specific differences in inflammatory markers at JIA diagnosis were restricted to the juvenile psoriatic arthritis (PsA) subtype, where females had statistically significantly higher ESR and CRP levels than males (median 26 [IQR 8-48.5] vs. 13 [7-21], p = 0.0169 and median 8 [IQR 4.5-13.5] vs. 5 [IQR 4-7], p=0.013), respectively. No significant sex-related differences were observed in other JIA subtypes (**Table 1 suppl**.)

### Impact of sex on timing to JIA treatment initiation and outcomes at 6 and 12 months

Time to methotrexate initiation was shorter in girls than in boys with JIA (log-rank p = 0.015, **Figure 2A**). In contrast, no sex difference was observed in time from baseline assessment when the diagnosis was made to biologic treatment initiation (log-rank p = 0.31, **Figure 2B**).

**Figure 2:**
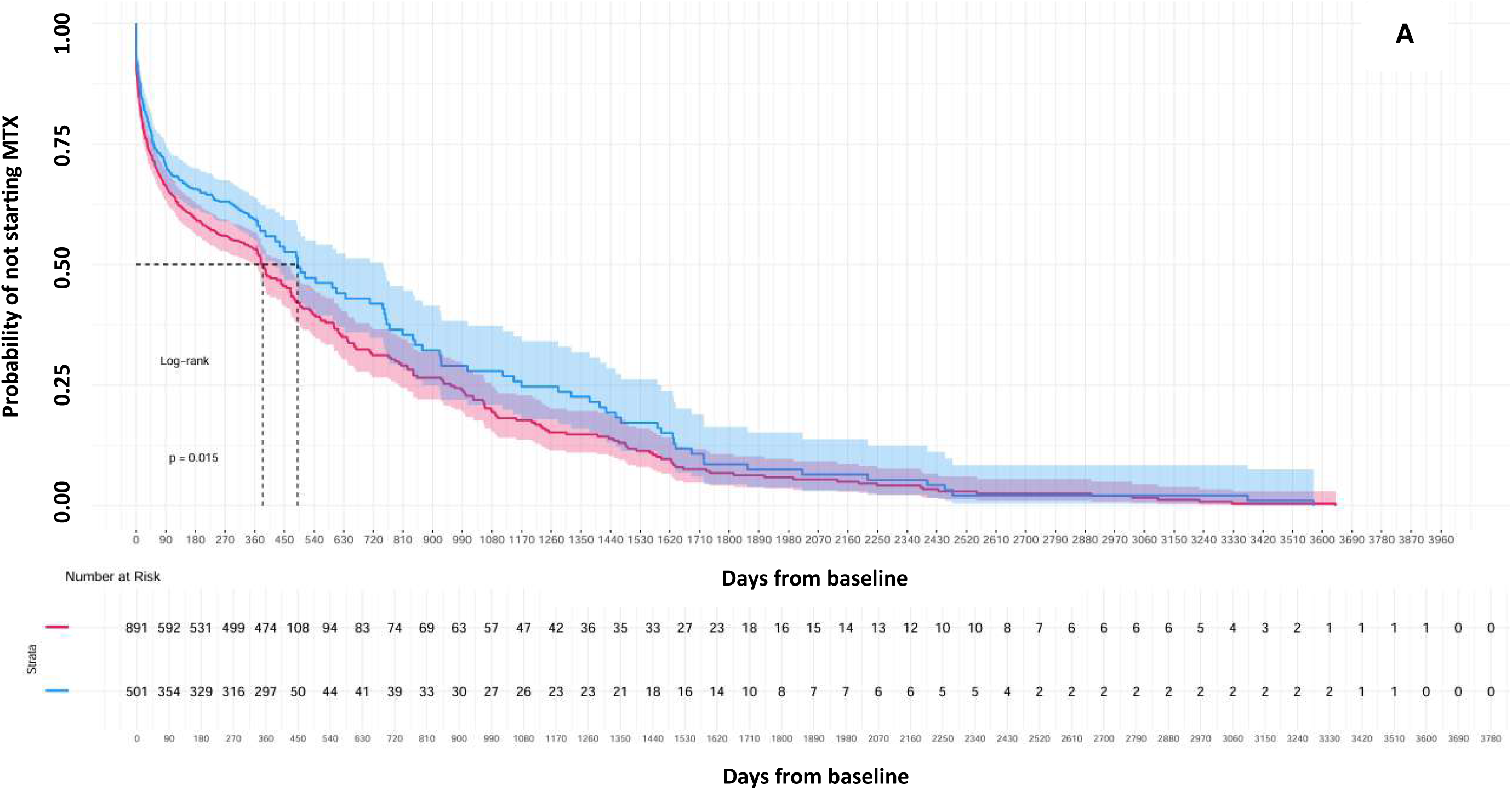
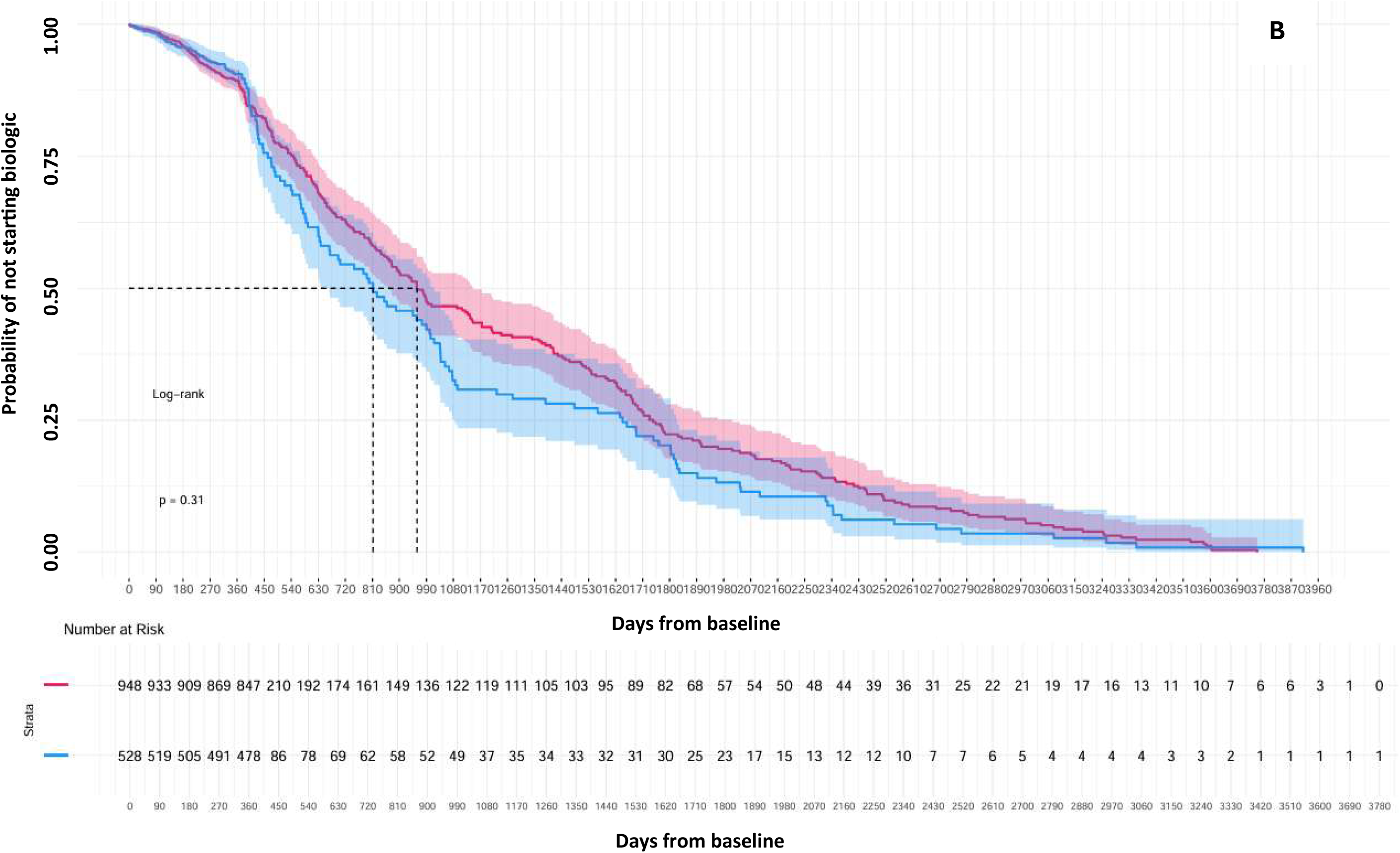
Time from baseline to treatment initiation. **A.** Time from baseline to methotrexate treatment initiation. **B.** Time from baseline to biologic treatment initiation. *Cox proportional hazards regression was used to compare time (in days) to methotrexate treatment initiation from JIA diagnosis between boys (In Blue) and girls (in Red). Differences between groups were assessed using the log-rank test. Legend; MTX- methotrexate*.

Cox regression analyses likewise showed no association between sex and time from baseline evaluation to either methotrexate or biologic initiation. Adjusted hazard ratios were 0.89 (95% CI 0.74-1.06, p = 0.21) for methotrexate initiation and 0.91 (95% CI 0.72-1.16, p = 0.47) for biologic initiation after accounting for age at onset, symptom duration, concomitant uveitis, and ILAR JIA category (**Figure 3A**).

**Figure 3:**
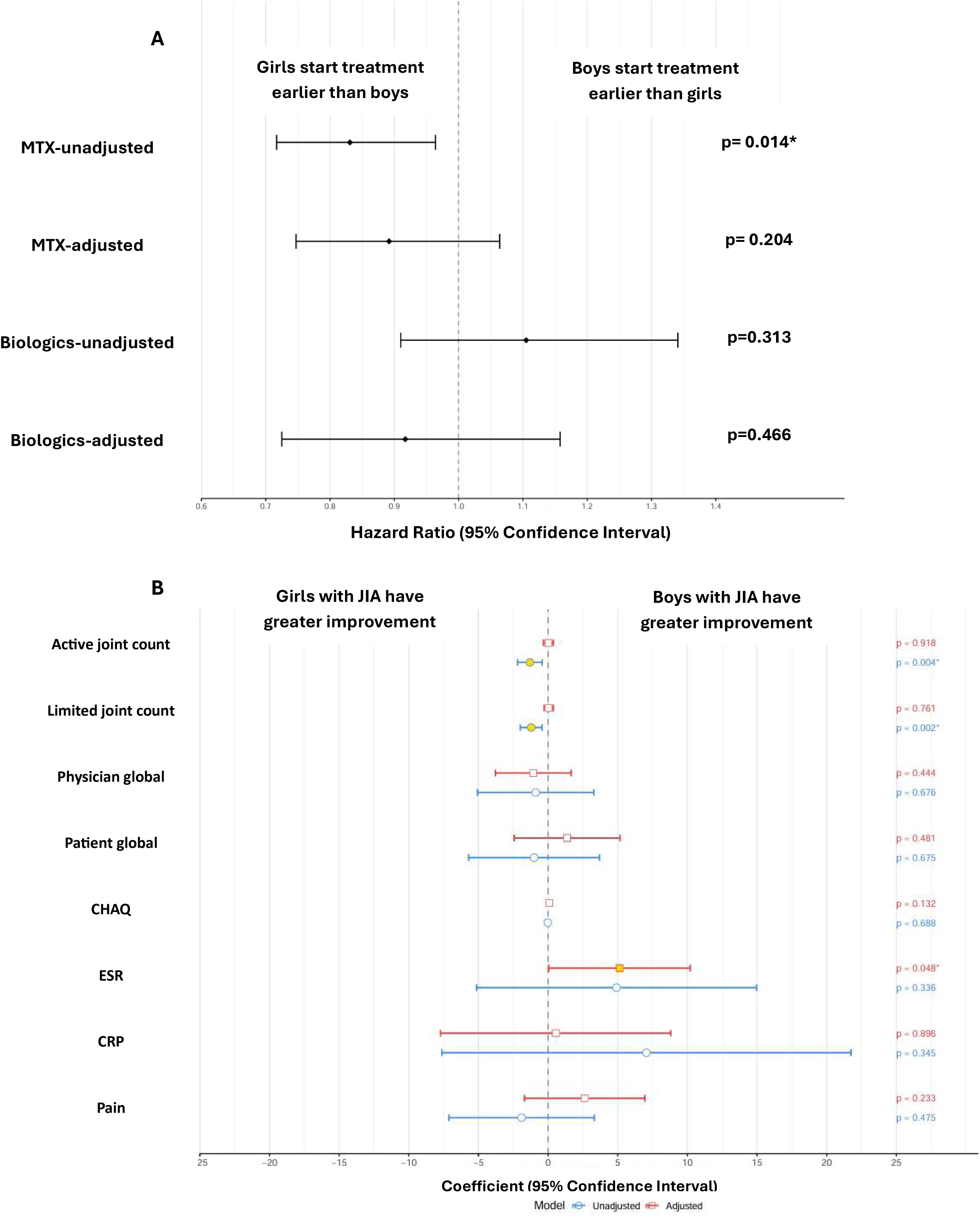
Forest plot of univariate and multivariate regression analyses comparing boys and girls with JIA. **A.** Forest plot showing hazard ratios for time to methotrexate and biologic treatment initiation for adjusted and non-adjusted regression analyses are shown with 95% confidence intervals. **B**. Forest plot showing regression coefficients for 12-month outcomes in boys compared with girls with JIA. *Estimates are presented with 95% confidence intervals. p<0.05 was considered significant*.

Interestingly, among CYP who had already received a JIA diagnosis before referral to paediatric rheumatology and entry into this inception cohort, the interval from diagnosis to treatment initiation was significantly shorter in girls than in boys (girls: n = 60, median 25 days [IQR 6.8-93.8] vs. boys: n = 28, median 61 days [IQR 13.8-179.2]; p = 0.0006).

Six-month follow-up outcomes showed no significant sex-based differences (Suppl. Table 2) (**Suppl. Table 2**). In unadjusted analyses, both active joint count (β = -1.30, 95% CI -2.19 to -0.42, p = 0.004) and limited joint count (β = -1.22, 95% CI -1.99 to -0.44, p = 0.002) improvements at 12 months compared to baseline were significantly lower in boys compared to girls. However, these associations were fully attenuated after adjustment for confounders (active joints: β = 0.02, 95% CI -0.34 to 0.38, p = 0.918; limited joints: β = 0.05, 95% CI -0.28 to 0.38, p = 0.761) (**Figure 3B**).

ESR was the only outcome showing a significant adjusted improvement in boys compared to girls at 12 months (β = 5.12, 95% CI 0.04 to 10.20, p = 0.048). All other outcomes—including physician global and parent global disease assessment (VAS 0-10), Childhood Health Assessment Questionnaire (CHAQ), CRP, and pain VAS—showed no significant associations in either unadjusted or adjusted models (all p>0.05, see **Figure 3B**).

## Discussion

This study, one of the largest contemporary JIA cohorts, provides a comprehensive assessment of sex-specific differences in disease onset, clinical course, and outcomes. Our findings confirm a non-uniform timeline of JIA manifestation across sexes. The marked female predominance (64.3% vs. 35.7%) aligns with established epidemiology (25, 26), and the younger age at symptom onset and diagnosis in girls suggests earlier disease expression, potentially reflecting sex-linked biological susceptibility windows influenced by genetic and hormonal factors.

The overall cohort demonstrated a multimodal age distribution, consistent with previous reports, but sex-stratified analyses revealed distinct patterns. Girls showed clear bimodality, echoing Swedish data (27, 28), whereas boys displayed a flatter distribution, likely shaped by the predominance of ERA. These findings highlight the need for sex-specific epidemiological models: bimodality in girls may reflect hormonally mediated susceptibility periods, while male patterns are weaker or confounded by subtype composition. This framework helps explain inconsistencies across studies, as cohorts with higher female representation are more likely to demonstrate bimodality, whereas differing sex or ethnic structures may obscure it.

In our cohort, the median delay from symptoms onset to baseline diagnosis was 4.4 months, substantially shorter than the 6.26 months (27.2 weeks [IQR 14.1-66 weeks]) delay reported in UK adults with rheumatoid arthritis (29). No sex differences were observed in diagnostic delay. The right-skewed distribution in both sexes suggests that systemic factors, such as referral pathways and access to paediatric rheumatology, affect boys and girls, within the UK’s relatively uniform healthcare structure. Importantly, the dedicated referral routes for childhood arthritis did not contribute to delay, in contrast to adults with RA, where longer delays are linked to referral from primary care to adult rheumatology.

Sex is firmly established as a fundamental determinant of JIA phenotype, with clear dimorphism in subtype distribution (30). Girls predominated in RF-negative and RF-positive polyarthritis, consistent with paediatric and adult patterns (27, 31, 32) and accounted for over two-thirds of oligoarthritis cases in our cohort, aligning with published data (33, 34).

Conversely, ERA showed a strong male predominance, as consistently described in international studies (35, 36). Its later onset, often around 11-12 years, and subtler axial or hip involvement likely contribute to diagnostic delays in boys (37).

In this cohort, at diagnosis, girls presented with a higher number of active joints than boys, whereas limited joint counts were comparable between the sexes. This pattern suggests girls may have exhibited a slightly greater degree of clinically apparent synovitis at onset, which likely reflects underlying differences in JIA subtype distribution rather than intrinsic sex-based differences in inflammatory severity. Girls are more frequently affected by early-onset, ANA-positive phenotypes associated with greater synovitis, whereas boys more often present with enthesitis-related or HLA-B27-associated disease, which typically involves fewer swollen joints (38).

Although uveitis prevalence did not differ significantly by sex in our overall cohort (females: 5.4%, males: 5.0%; p = 0.916), this likely reflects limited case numbers and the counterbalancing effect of male-dominant ERA, itself linked to uveitis. Crucially, equal uveitis prevalence, although reported in other cohorts (39) does not imply equal burden, as males with ERA often experience more severe disease trajectories (14). Stratification by sex and ILAR subtype is therefore essential to disentangle risk, refine prognostic models, and capture the nuanced ways in which sex shapes susceptibility and long-term outcomes in JIA.

The absence of sex differences in uveitis prevalence overall, despite higher active joint counts in girls, supports the view that sex effects in JIA are phenotype-specific rather than uniform across the disease spectrum. Across most ILAR subtypes, acute-phase reactants did not differ by sex, indicating that systemic inflammatory burden is driven primarily by subtype rather than sex. This is notable given well-described sex-dimorphic immune responses, with females typically exhibiting stronger innate and adaptive activation (40) and documented effects of sex and puberty on paediatric rheumatic disease outcomes (41, 42). In our cohort, inflammatory burden aligned closely with ILAR subtype: median CRP levels were highest in systemic JIA (90-139 mg/L), modest in polyarticular JIA, and minimal in ERA (5-10 mg/L).

Sex-divergent elevations in inflammatory markers were observed only in the PsA subtype. This should be interpreted cautiously given the small sample size and lack of adjustment for BMI or other confounders. Unlike adult PsA, where CRP and ESR do not consistently differ by sex (43), girls with juvenile PsA in our cohort had significantly higher inflammatory markers than boys. This apparent reversal of the adult pattern likely reflects the clinical heterogeneity of juvenile PsA, which comprises two phenotypes: an early-onset, ANA-positive, female-predominant form resembling oligoarticular/polyarticular JIA, and a later-onset, male-predominant form with enthesitis, axial disease, and strong HLA-B27 association (44). Our cohort appears enriched for the early-onset phenotype, consistent with the female-to-male ratio of 1.57:1. A recent systematic review of adult PsA similarly reported greater overall disease burden in women, more peripheral involvement and higher tender joint counts, despite no sex differences in CRP (43).

When interpreted together, these findings suggest that sex differences in inflammatory markers in juvenile PsA likely reflect phenotype distribution rather than uniform sex-based immune reactivity. Nonetheless, subgroup results should be interpreted cautiously given limited power and residual confounding.

Girls appeared to start methotrexate earlier than boys in unadjusted analyses, suggesting a sex difference in early treatment escalation; however, no sex difference was seen in time to biologic initiation. After adjusting for age at onset, symptom duration, uveitis, and ILAR category, sex was not independently associated with time to either methotrexate or biologic therapy. This indicates that earlier methotrexate use in girls likely reflects underlying disease characteristics rather than sex itself, and that treatment escalation pathways are comparable once clinical factors are considered. Unlike adult rheumatology, where women often experience delays (45, 46), paediatric care in this UK cohort showed equitable access, likely reflecting NHS standardised protocols, centralised referral pathways, and universal healthcare. A non-significant trend toward earlier biologic use in boys was attributable to the higher prevalence of ERA, where biologics are introduced earlier, rather than systemic bias. Overall, these findings support a sex-neutral, disease-driven approach consistent with treat-to-target principles, while underscoring the importance of monitoring equity in less standardised health-care systems

In unadjusted analyses, boys showed smaller 12-month improvements in active and limited joint counts, but these differences disappeared after adjustment for age, ILAR subtype, joint count and presence of uveitis at baseline, indicating that crude associations reflected underlying disease rather than sex. The only adjusted sex-specific signal was a slightly greater ESR improvement in boys. All other clinical and patient-reported outcomes, including global assessments, CHAQ, and pain, showed no meaningful sex differences in either unadjusted or adjusted models. Similarly, a recent population-based study found that typical sex differences in depression were not seen in JIA, although CYP with JIA, particularly those with public insurance, had higher anxiety rates. These findings reinforce the need for integrated JIA care that addresses both physical and mental health (47).

Overall, in this study, sex did not appear to be a major determinant of early treatment response, and apparent disparities were largely driven by baseline features and subtype-related pathways.

Our findings contrast with a recent analysis of 9,601 CYP with JIA, which reported strong demographic gradients in disease burden: higher pain, global assessments, and JADAS10 scores with older age; greater disease activity in non-Hispanic Black compared with non-Hispanic White individuals; and consistently higher scores in females (48). This divergence likely reflects differences in study design and healthcare context. Harris et al. assessed cross-sectional burden, whereas we examined change over the first 12 months and adjusted for baseline activity and subtype, key drivers of sex differences. The cohorts also sit within very different health systems: the NHS provides universal, standardised access to paediatric rheumatology and biologics, while the U.S. system is shaped by fragmented insurance coverage and marked racial and socioeconomic inequities. Such structural factors may amplify demographic disparities in U.S. cohorts and attenuate them in settings with more uniform access.

Although detailed sex-specific mechanisms lie beyond the scope of this analysis, existing evidence indicates that sexual dimorphism in JIA arises from distinct immunological pathways, including strong associations with polygenic risk scores (49), and pronounced female-biased B-cell dysregulation in synovial fluid in oligoarthritis (50), underscoring the complex ways in which sex shapes disease biology and outcomes in JIA.

This study’s multicentre design across seven UK paediatric rheumatology centres, its prospective inception cohort, and its large sample size provide strong generalisability and statistical power, with standardised NHS care further reducing variability. A major strength is the explicit evaluation of sex as an independent biological factor, supported by rigorous adjustment for confounders.

Limitations include the absence of adjustment for RF/anti-CCP, HLA-B27, and BMI; relatively short follow-up; and potential bias from missing data without imputation. Analyses of smaller subtypes and inflammatory markers were underpowered. Nonetheless, the study design and available adjustments support confidence in the main findings. Sex was recorded as assigned at birth, with no information on intersex status or gender identity, underscoring the need for future work incorporating these dimensions.

Sex appears to influence JIA mainly through its association with subtype and baseline phenotype rather than directly shaping inflammation or treatment timing, though the female-predominant burden in PsA warrants further study. These findings support a personalised medicine approach that integrates sex with immunopathology and clinical presentation. Longer follow-up, broader confounder adjustment, and replication in other health systems will determine how generalisable the equitable treatment patterns seen in the UK are. Overall, precision strategies incorporating sex, subtype, and biomarker profiles may refine care further, while our data suggest no major sex-based differences in early outcomes.

## Data Availability

The data and analytic code used in this study will be made available upon reasonable request to the CAPS study team, following approval from the University of Manchester.

