## Supplementary material for "Investigating the Impact of Sex on Outcomes in Juvenile Idiopathic Arthritis"

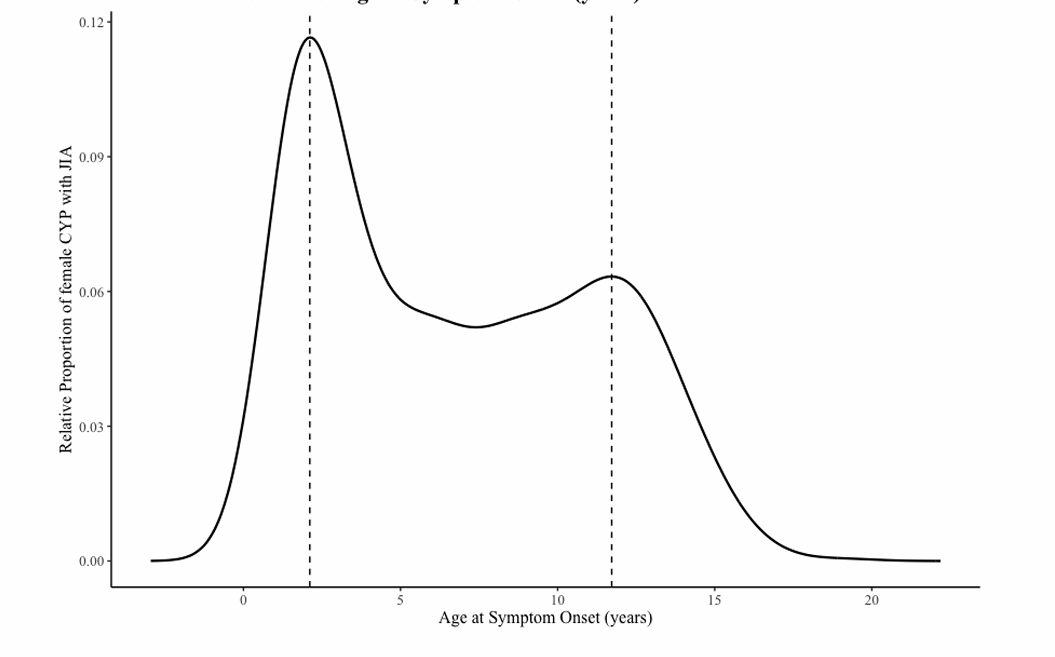

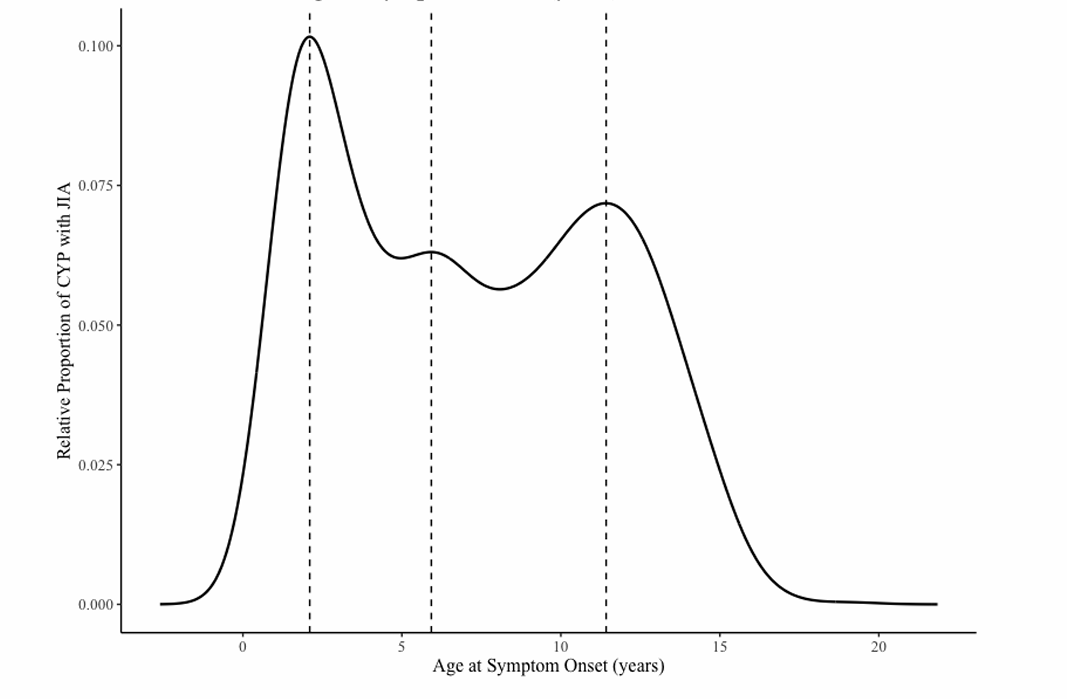

**B**
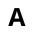

**A**

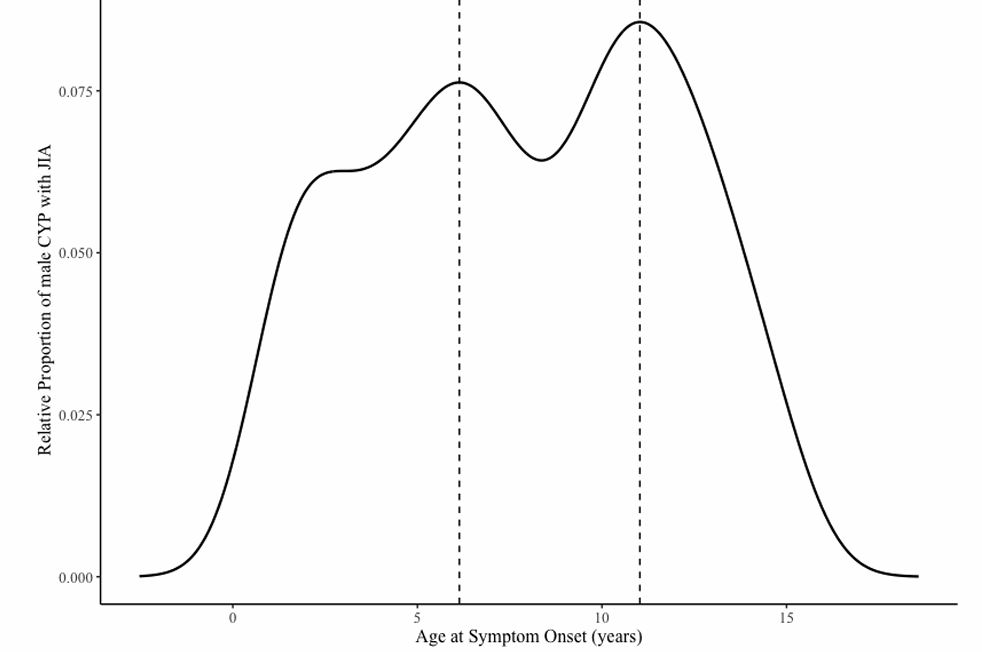

**C**

**Suppl. Figure 1. Distribution of age at symptoms onset in JIA.** A. The distribution of age at symptoms onset in the entire JIA cohort. B. The distribution of age at symptoms onset in females. C. The distribution of age at symptoms onset in males. *Kernel density curve with individual ages (in years) marked by vertical lines.*

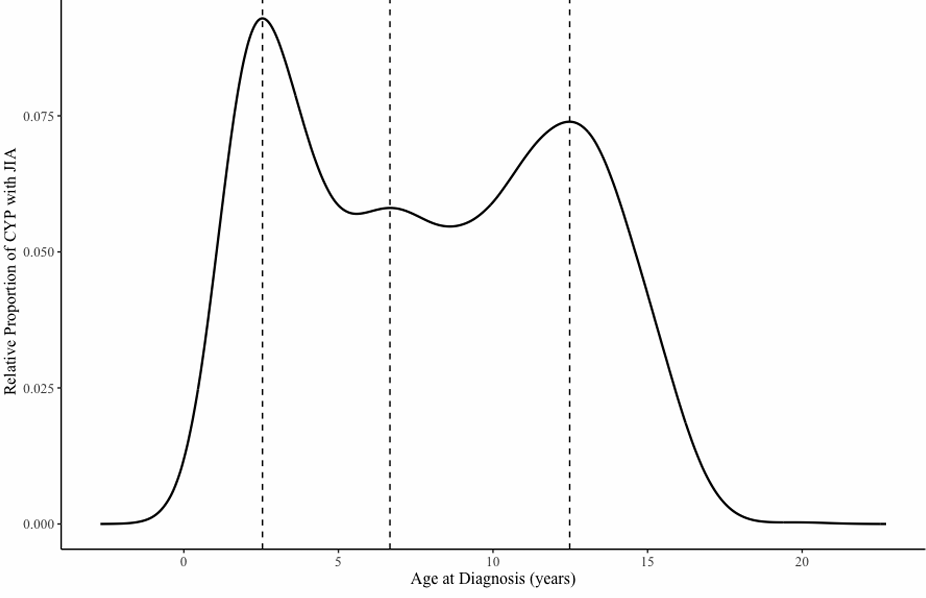

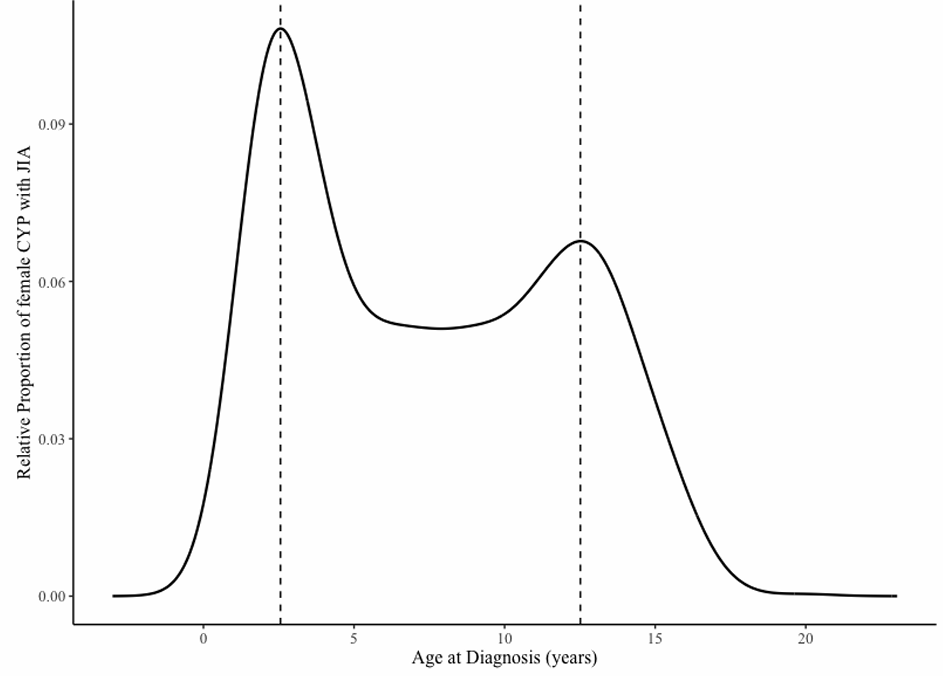

**A**

**B**
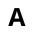

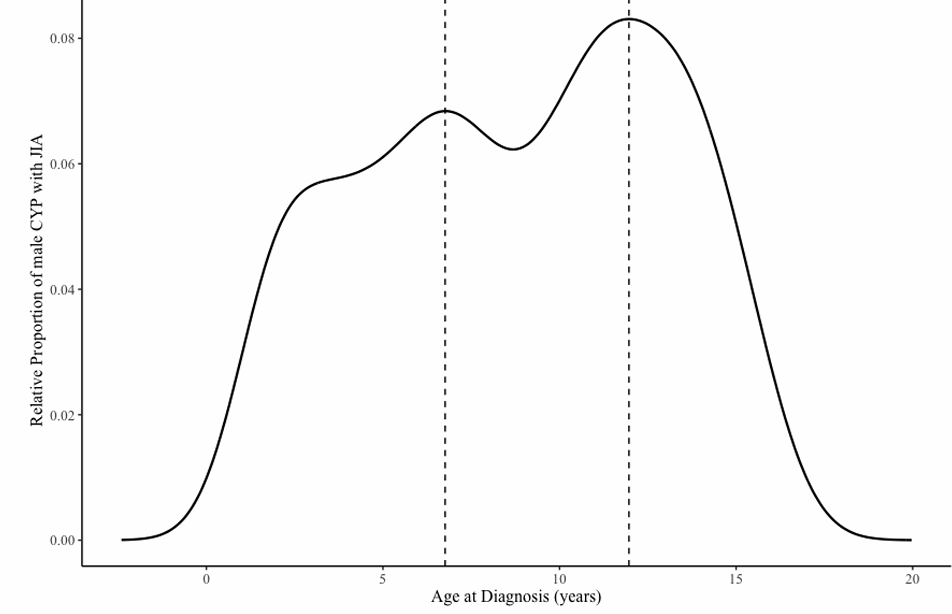

**C**

**Suppl. Figure 2. Distribution of age at JIA dignosis.** A. The distribution of age at JIA diagnosis in the entire JIA cohort. B. The distribution of age at JIA diagnosis in females. C. The distribution of age at at JIA diagnosis in males. *Kernel density curve with individual ages (in years) marked by vertical lines.*

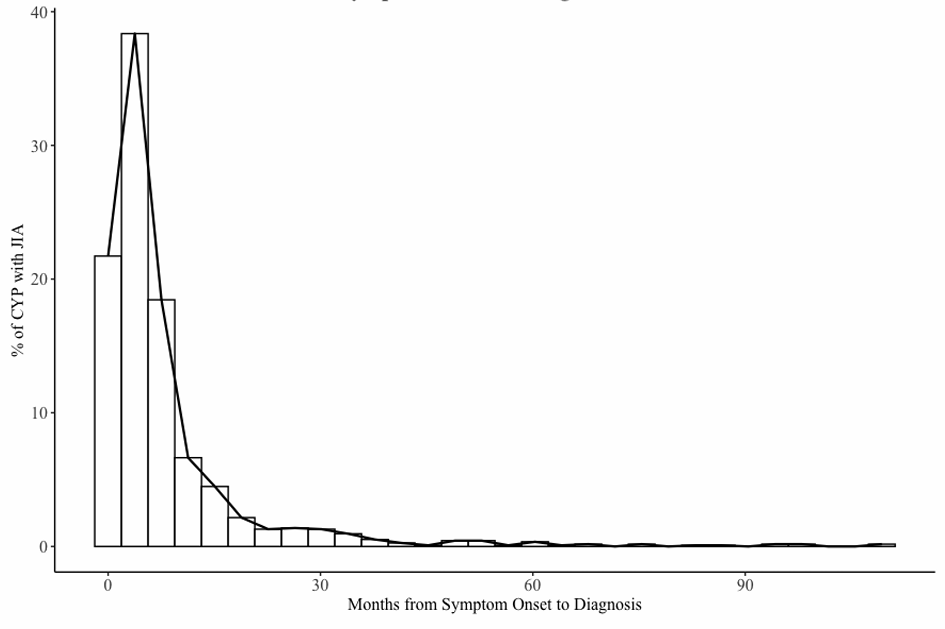

**A**

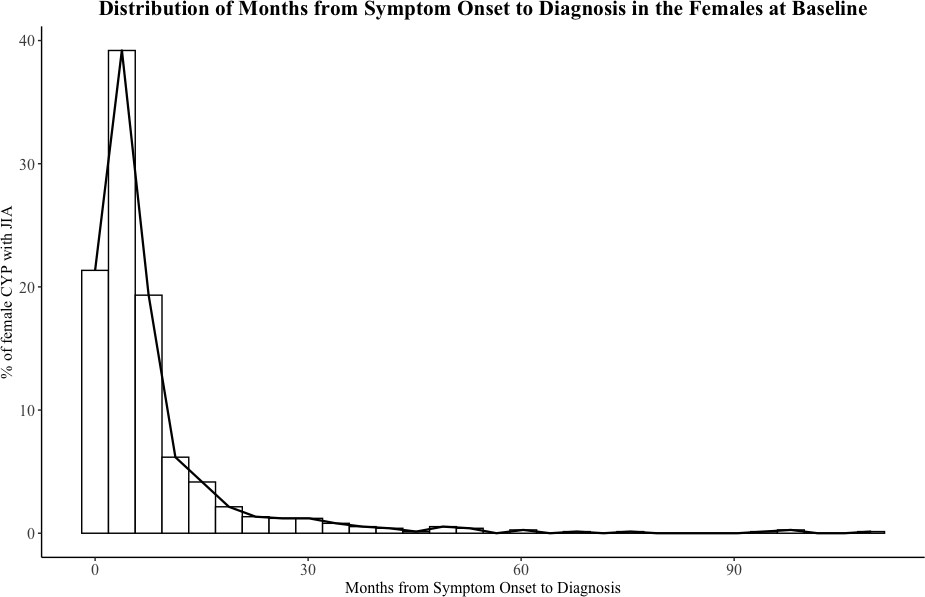

**B**

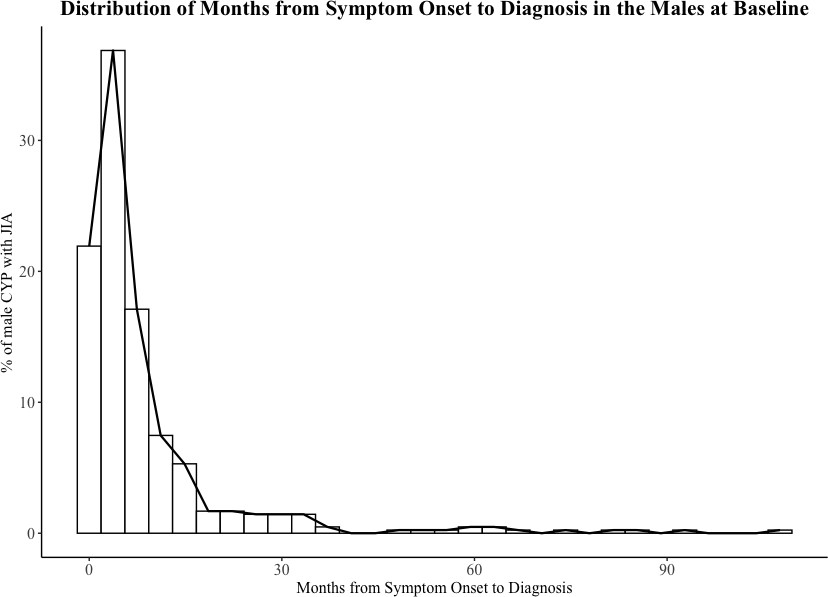

**C**

**Suppl. Figure 3. Distribution of delay in JIA diagnosis (as recorded at baseline).** A. For the entire JIA cohort. B. For females. C For males. *Histogram illustrating diagnostic delay in months. The boxed bars indicate how many patients fall into each delay category.*

**Table 1 Supplementary.** Inflammatory markers at baseline (diagnosis) stratified by sex and JIA ILAR category

| **ILAR subtype** | **ESR (mm/h)**  **median (IQR) (n)** | ***P*-value** | **CRP (mg/L)**  **median (IQR) (n)** | ***P*-value** |
| --- | --- | --- | --- | --- |
| **sJIA** |  | 0.414 |  | 0.0614 |
| Female  Male | 79 (60, 98) (n =33)  86.5 (67.8, 109.8) (n = 20) |  | 90.6 (52.2, 146.5) (n = 34)  139.3 (89.2, 245.2) (n = 22) |  |
| **Oligo-persistent** |  | 0.598 |  | 0.101 |
| Female  Male | 17 (6.5, 33.5) (n = 167)  19 (6, 40) (n = 85) |  | 5 (4, 9) (n = 153)  7 (4, 18.2) (n = 80) |  |
| **Oligo- extended** |  | 0.444 |  | 0.217 |
| Female  Male | 35 (19, 43) (n = 13)  21.5 (13.2, 29.8) (n = 2) |  | 8 (5.5, 15.5) (n = 14)  5 (5, 5.5) (n = 4) |  |
| **RF- poly JIA** |  | 0.847 |  | 0.96 |
| Female  Male | 32 (13.8, 58.2) (n = 100)  25 (6.5, 62.2) (n =28) |  | 14 (6, 33) (n = 89)  16.2 (6, 32.1) (n = 26) |  |
| **RF+ poly JIA** |  | NA |  | 0.615 |
| Female  Male | 55 (14, 102.5) (n = 19)  20 (20, 20) (n = 1) |  | 19 (10, 47.2) (n = 18)  14 (9.5, 14) (n = 3) |  |
| **ERA** |  | 0.736 |  | 0.133 |
| Female  Male | 33.5 (28.5, 39.2) (n = 6)  30 (19.8, 51.5) (n = 24) |  | 5 (4, 5.6) (n = 5)  10 (7, 25.5) (n = 23) |  |
| **PsA** |  | **0.0169** |  | **0.013** |
| Female  Male | 26 (8, 48.5) (n = 27)  8 (4.5, 13.5) (n = 19) |  | 13 (7, 21) (n = 25)  5 (4, 7) (n = 18) |  |
| **UA** |  | 0.2204 |  | 0.614 |
| Female  Male | 23 (9, 46) (n = 85)  20 (5, 41.5) (n = 51) |  | 8.8 (4, 31.8) (n = 78)  7 (4, 25) (n = 49) |  |

***Legend:*** *CRP - C‑reactive protein; ERA - Enthesitis‑related arthritis; ESR - Erythrocyte sedimentation rate; Oligo‑extended - Extended oligoarticular JIA; Oligo‑persistent - Persistent oligoarticular JIA; PsA - Psoriatic arthritis; RF‑ polyJIA - Rheumatoid factor–negative polyarticular JIA; RF+ polyJIA - Rheumatoid factor–positive polyarticular JIA; sJIA - Systemic onset JIA; UA - Undifferentiated arthritis*

**Table 2 Supplementary.** Sex‑stratified early outcomes at 6‑month follow‑up in JIA

| **Outcomes** | **Data availability within**  **the whole cohort, n (%)** | **n (%) or**  **median**  **(IQR)** | **Female** | **Female Entries**  **Available** | **Male** | **Male Entries**  **Available** | **P value** |
| --- | --- | --- | --- | --- | --- | --- | --- |
| Active joint count | 894 (82.3%) | 0 (0, 2) | 0 (0, 2) | 575 | 0 (0, 1) | 319 | 0.342 |
| Limited joint count | 894 (82.3%) | 0 (0, 2) | 0 (0, 2) | 575 | 0 (0, 1) | 319 | 0.561 |
| Presence of uveitis | 859 (79.1%) | 44 (5.12%) | 31 (5.7%) | 547 | 13 (4.2%) | 312 | 0.334 |
| CHAQ score | 652 (60.0%) | 0.25  (0.00, 1.00) | 0.25  (0.00, 1.00) | 432 | 0.25  (0, 0.906) | 220 | 0.892 |
| Parent global  assessment* | 617 (56.8%) | 7 (1, 31) | 8 (1, 28) | 412 | 6 (1, 37) | 205 | 0.123 |
| Physical global  assessment* | 743 (68.4%) | 8 (1, 21) | 8 (1, 21.25) | 476 | 7 (1, 21) | 267 | 0.811 |

*measured on a visual analogue scale 0-100

***Legend****: CHAQ - Childhood Health Assessment Questionnaire.*
